# Injuries and fatalities in Colombian mining emergencies (2005 - 2018)

**DOI:** 10.1101/2021.04.04.21254888

**Authors:** Gloria C. Gheorghe, Edgar F. Manrique-Hernández, Alvaro J. Idrovo

**Affiliations:** Safety and Rescue Mining Group, Agencia Nacional de Minería. Bogotá DC, Colombia; Safety and Health in Work Program, Medicine and Health Sciences School, Universidad del Rosario. Bogotá DC, Colombia; Public Health Department, School of Medicine, Universidad Industrial de Santander. Bucaramanga, Colombia

**Keywords:** occupational health, injuries, mortality, mining, Colombia

## Abstract

**Background:** During the last decades several developed countries reported a decrease in the occurrence of mining injuries. Mining is a very important sector of Colombia’s economy without analyses of injuries and fatalities in mining emergencies.

**Objective:** This study describes the occurrence of mining emergencies in Colombia between 2005 and 2018, and their principal characteristics.

**Methods:** An ecological study was performed with the mining emergencies registered by the National Mining Agency between 2005 and 2018. The study described the place of occurrence, type of event, legal status and type of mines, mineral extracted, and number of injuries and fatalities. Benford’s law was used to explore the quality of the data.

**Results:** A total of 1,235 emergencies occurred, with 751 injured workers and 1,364 fatalities. The majority of emergencies were from collapses, polluted air, and explosions, most of which occurred in coal (77.41%), gold (18.06%), and emerald (1.38%) mines. Many emergencies occurred in illegal mines (27.21%), most of which were gold, construction materials, emerald, and coal. Illegal mines had a higher relative proportion of injuries and fatalities than legal mines (*p<*0.05). Mining disasters are likely to be underreported given that Benford’s Law was not satisfied.

**Conclusions:** Colombia is a country with increasing mining activity, where the occurrence of mining emergencies, injuries, and fatalities is growing. This is the first one full description of mining emergencies in Colombia with the few available data.

## INTRODUCTION

Although injuries and fatalities are a frequent occurrence in the mining industry ^1^, several developed countries have seen a decrease in the occurrence of mining injuries over recent decades. Some of the best examples include several countries in the European Union as well as Canada and the United States ^2^. In contrast, injuries and fatalities in low- and middle-income countries have mostly occurred in informal high-hazard work with unsafe environments ^3^, such as a variety of mining activities. In addition, historical analyses indicate that large open-pit mining has improved working conditions, while the environmental sustainability of mining has decreased ^4^.

Mining has become a key income-generating industry in Colombia over recent decades, surpassing agricultural activities and industries that had previously been dominant ^5^. The largest operations involve the extraction of coal, ferronickel, and gold, followed by building materials, salt, silver, and platinum ^6^. In spite of the growing importance of mining to Colombia’s economy, occupational health and environmental studies of the mining sector are scarce. In 2018, roughly 12% of miners were injured, and the rate of fatality was 73 cases per 100,000 workers, with the mining sector having the largest number of deaths ^7^. Given this context, the objective of the present study was to describe the occurrence of injuries and fatalities caused by mining emergencies from 2005 to 2018.

## MATERIALS AND METHODS

An ecological study was performed with mining emergencies in Colombia as the unit of analysis ^8^. The study reviewed emergency statistics that were registered between 2005 and 2018 by the National Mining Agency (NMA, Agencia Nacional Minera). The NMA is the mining authority in Colombia, whose functions include: granting mining contracts to private entities, managing mining land registries, monitoring mining owners’ compliance with contractual obligations, including technical, legal, and financial obligations (royalties payments and other financial compensations), mining health and safety, and coordinating the National Mining Rescue System. Thus, the NMA registers and is responsible for only the most serious mining emergencies.

The variables included in the analysis were date of occurrence of the emergency, mineral extracted, location (department, municipality, and town), type of mine (open-pit or underground), legal status of the mine (legal or illegal), type of event (falling from another level, collapse, electrical, mechanical, explosion, slope instability, fire, flood, polluted air, or events involving heavy machinery), and people affected (injuries and deaths).

The statistical methods consisted of using percentages to describe the categorical variables and measures of central tendency and dispersion to describe quantitative variables, based on evaluating the distribution with the Shapiro-Wilk test. The Mann-Whitney test was used to explore possible differences in the occurrence of injuries and fatalities in legal versus illegal mines. In addition, the quality of the data was evaluated by applying Benford’s Law (for first and second digits) to the injury and fatality data. For the analysis of first digits, data with zeroes were excluded ^9,10^. The *digdis* macro developed by Ben Jann (ETH Zurich) was used for that purpose, and Pearson’s *x*^*2*^ and log likelihood ratio were used to evaluate the goodness-of-fit of the distributions. The Stata 14 statistical program (Stata Corporation, College Station, TX, USA) was used for these analyses.

## RESULTS

A total of 1,235 mining emergencies occurred in Colombia between 2005 and 2018, with 751 injured workers and 1,364 fatalities. Thus, a mining worker was injured approximately every 6.8 days, and a miner died from a mining emergency every 3.75 days. Figure 1 lists the sites where the mining emergencies occurred. As can be seen, the municipalities with the most number of mining emergencies were Cucunuba (n=68), Lenguazaque (n=61), and Guacheta (n=52) in Cundinamarca; Marmato (n=55) in Caldas; Amagá (n=49) and Angelópolis (n=40) in Antioquia; and Tasco (n=41) in Boyacá.

**Figure 1.**
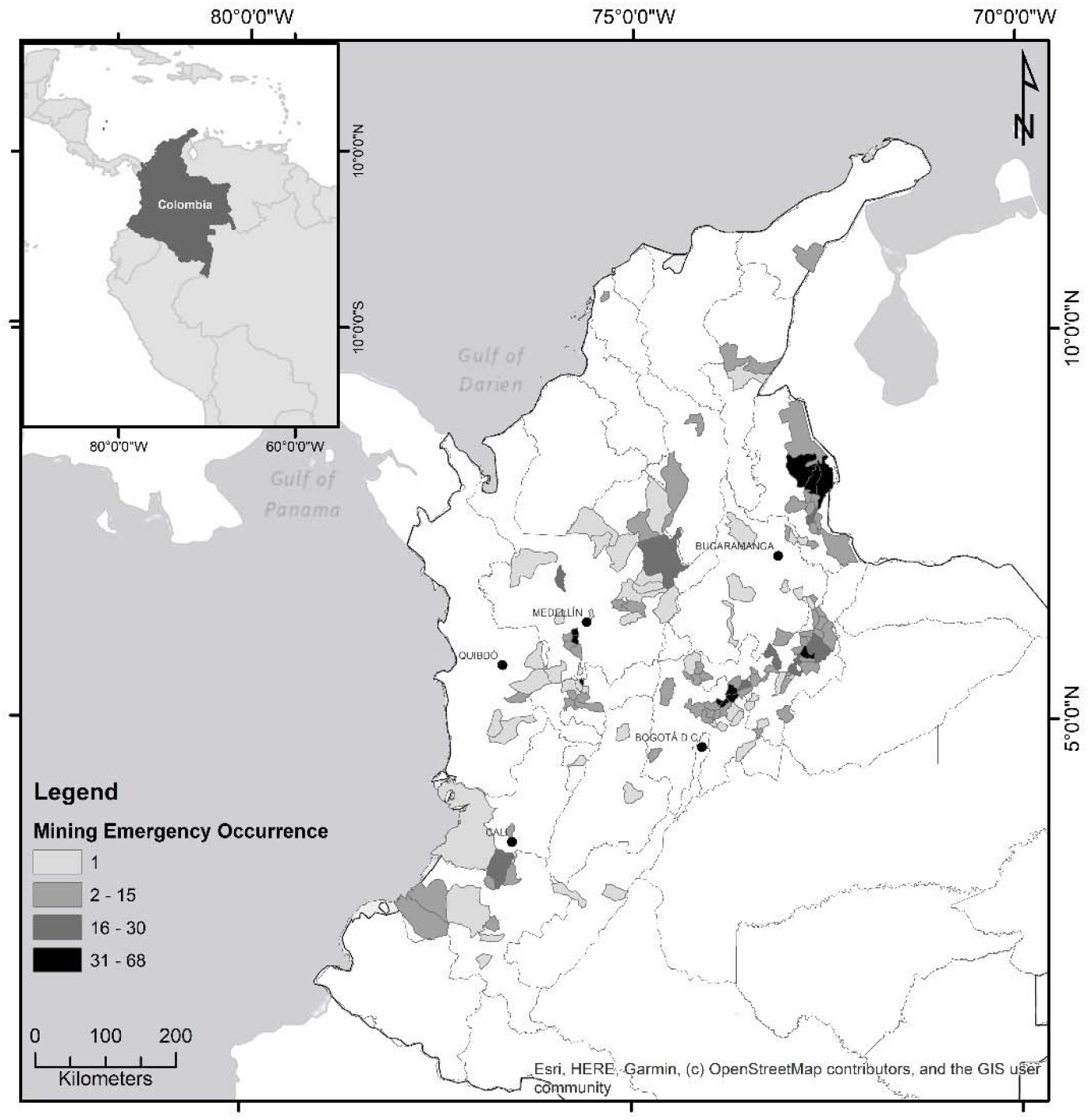
Map of sites with mining emergencies in Colombia (2005 - 2018)

Figure 2 presents the occurrence of mining emergencies by month, which shows large monthly variability in the number of emergencies (median 7, min 0, max 20), injuries (median 3, min 0, max 34), and fatalities (median 7, min 0, max 78). Table 1 presents the evaluation of the data quality, which shows a deviation from Benford’s Law of first digits. An evaluation of the second digits resulted in a similar finding (data not shown), which suggests an underreporting of workers affected by mining emergencies.

**Table 1.** Expected and observed frequencies of digits based on Benford’s law for the occurrence of injuries and fatalities in Colombian mining emergencies (2005-2018).*

| Digit | Expected (%) | Observed |  |  |  |
| --- | --- | --- | --- | --- | --- |
|  |  | Injuries (%) | P value | Fatalities (%) | P value |
| 1 | 30.103 | 58.228 | <0.0001 | 77.966 | <0.0001 |
| 2 | 17.609 | 22.152 | 0.3840 | 13.559 | 0.0019 |
| 3 | 12.494 | 10.127 | 0.2332 | 4.479 | <0.0001 |
| 4 | 9.691 | 4.747 | 0.0016 | 1.211 | <0.0001 |
| 5 | 7.918 | 1.266 | <0.0001 | 0.847 | <0.0001 |
| 6 | 6.695 | 1.266 | <0.0001 | 0.605 | <0.0001 |
| 7 | 5.799 | 1.582 | 0.0004 | 0.605 | <0.0001 |
| 8 | 5.115 | 0.316 | <0.0001 | 0.484 | <0.0001 |
| 9 | 4.576 | 0.316 | <0.0001 | 0.2 42 | <0.0001 |
| Pearson's $X^2$ | | $p<0.0001$ | | $p<0.0001$ | |
| Log likelihood ratio | | $p<0.0001$ | | $p<0.0001$ | |

\* mining emergencies with zero occurrences were excluded.

**Figure 2.**
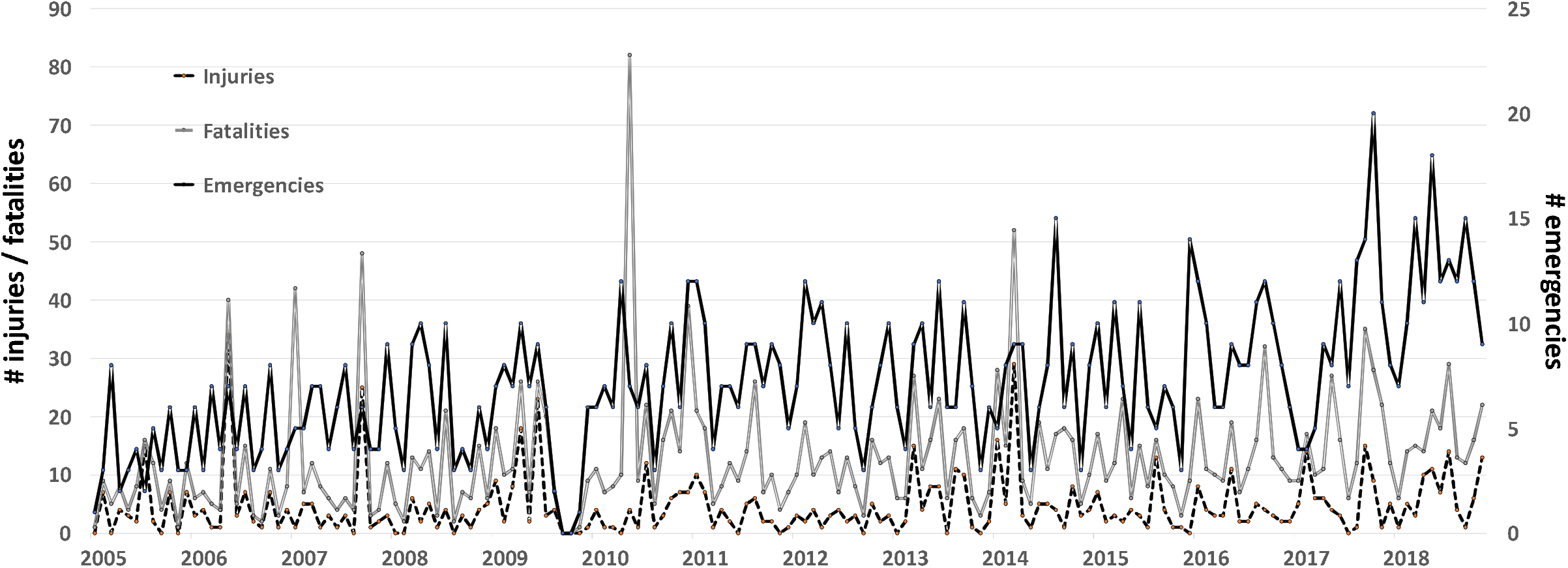
Temporal trend of injuries and fatalities in mining emergencies in Colombia (2005 – 2018)

Table 2 summarizes the main characteristics of the mining emergencies in Colombia between 2005 and 2018, most of which had few injured workers (97.81%). Fatalities had a similar distribution, with the majority of the emergencies having less than 5 deaths (97.49%). Most of the emergencies were due to collapses, polluted air, and explosions, which together represented 59.59% of total emergencies. The largest number of emergencies occurred on Wednesdays and the least number on Saturdays and Sundays, two days that typically are not workdays in Colombia. The mines where the emergencies occurred were coal, gold, and emerald, which together represented 96.85% of the total.

**Table 2.**
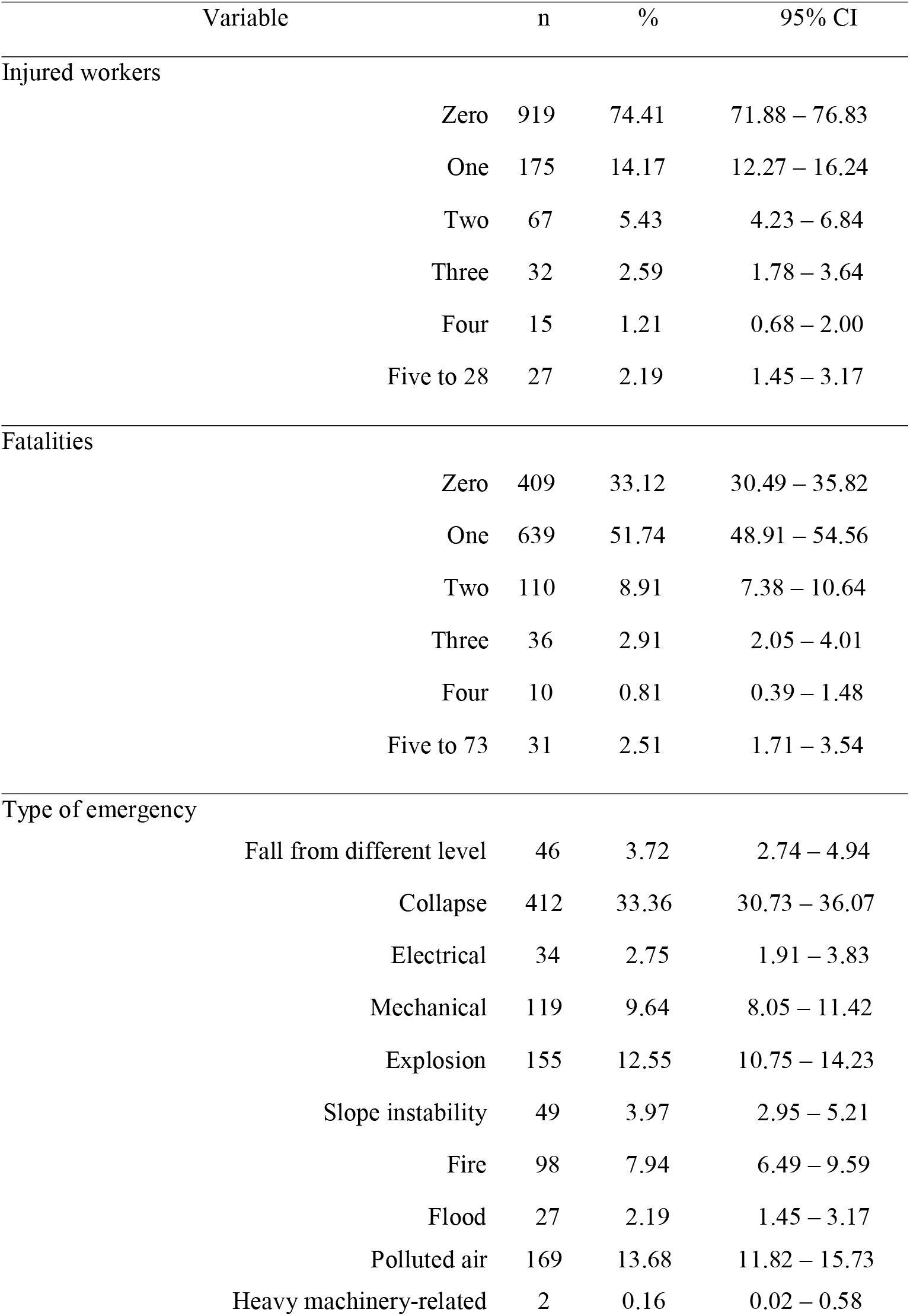
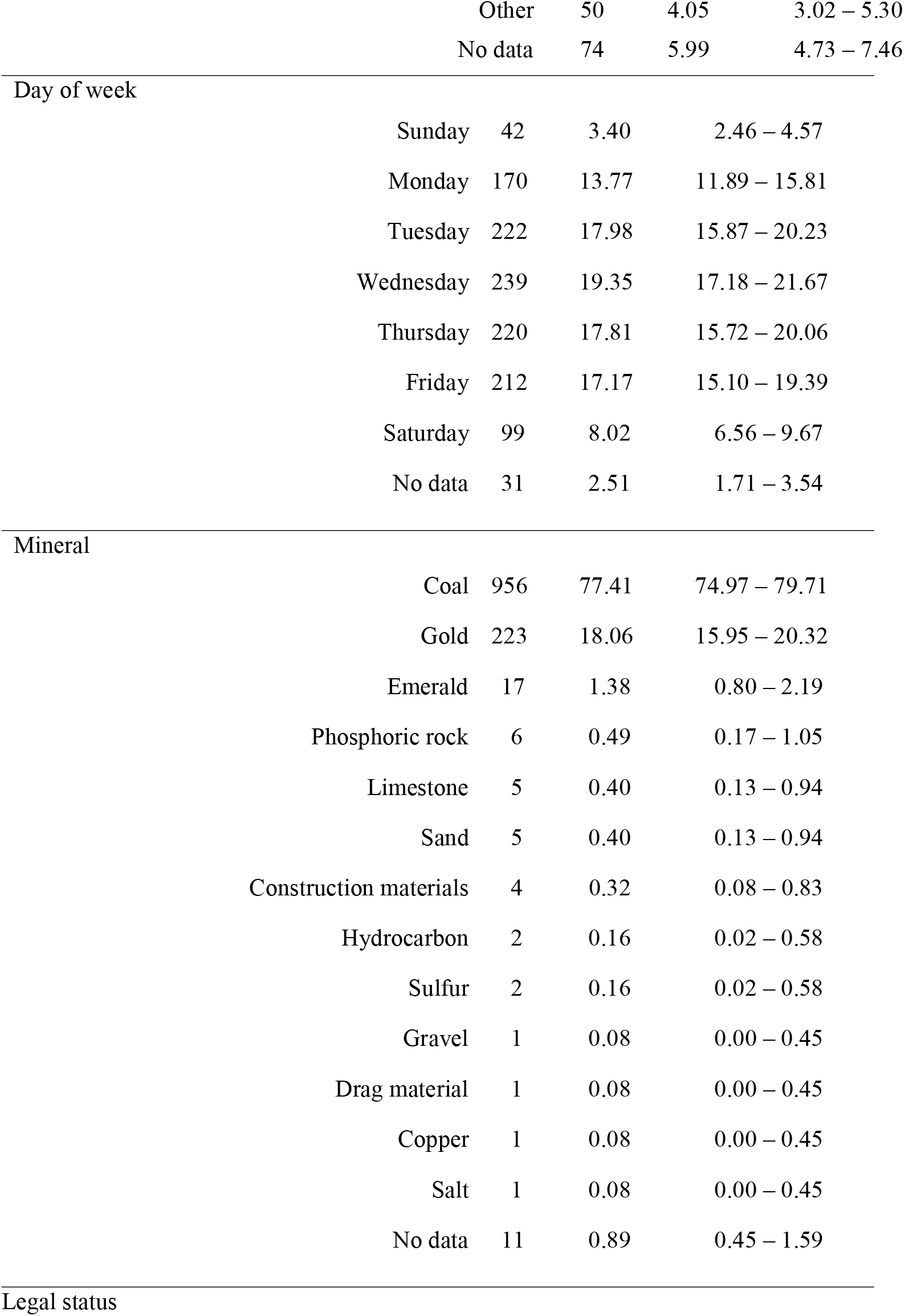
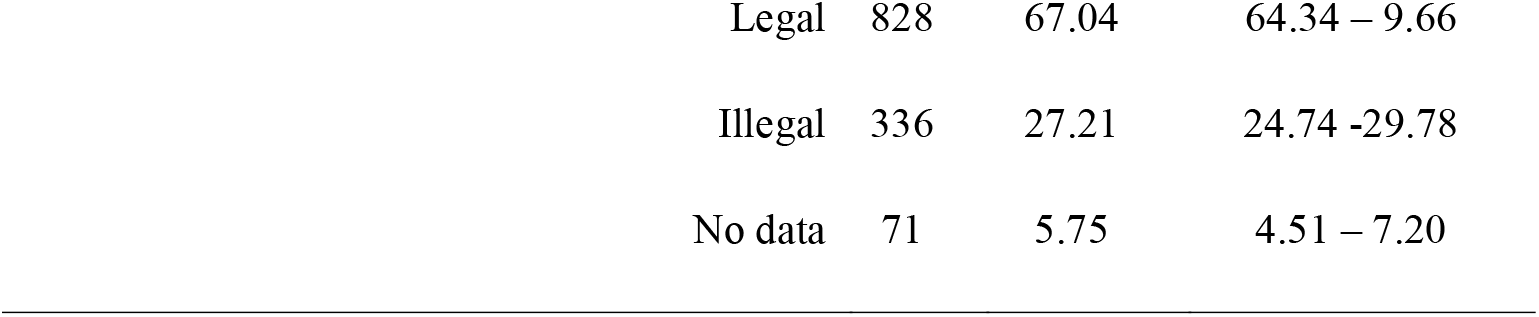
Characteristics of mining emergencies in Colombia (2005-2018)

| Variable | n | % | 95% CI |
| --- | --- | --- | --- |
| Injured workers |  |  |  |
| Zero | 919 | 74.41 | 71.88 – 76.83 |
| One | 175 | 14.17 | 12.27 – 16.24 |
| Two | 67 | 5.43 | 4.23 – 6.84 |
| Three | 32 | 2.59 | 1.78 – 3.64 |
| Four | 15 | 1.21 | 0.68 – 2.00 |
| Five to 28 | 27 | 2.19 | 1.45 – 3.17 |
| Fatalities |  |  |  |
| Zero | 409 | 33.12 | 30.49 – 35.82 |
| One | 639 | 51.74 | 48.91 – 54.56 |
| Two | 110 | 8.91 | 7.38 – 10.64 |
| Three | 36 | 2.91 | 2.05 – 4.01 |
| Four | 10 | 0.81 | 0.39 – 1.48 |
| Five to 73 | 31 | 2.51 | 1.71 – 3.54 |
| Type of emergency |  |  |  |
| Fall from different level | 46 | 3.72 | 2.74 – 4.94 |
| Collapse | 412 | 33.36 | 30.73 – 36.07 |
| Electrical | 34 | 2.75 | 1.91 – 3.83 |
| Mechanical | 119 | 9.64 | 8.05 – 11.42 |
| Explosion | 155 | 12.55 | 10.75 – 14.23 |
| Slope instability | 49 | 3.97 | 2.95 – 5.21 |
| Fire | 98 | 7.94 | 6.49 – 9.59 |
| Flood | 27 | 2.19 | 1.45 – 3.17 |
| Polluted air | 169 | 13.68 | 11.82 – 15.73 |
| Heavy machinery-related | 2 | 0.16 | 0.02 – 0.58 |

|  |  |  |  |  |
| --- | --- | --- | --- | --- |
|  | Other | 50 | 4.05 | 3.02 – 5.30 |
|  | No data | 74 | 5.99 | 4.73 – 7.46 |
| Day of week |  |  |  |  |
|  | Sunday | 42 | 3.40 | 2.46 – 4.57 |
|  | Monday | 170 | 13.77 | 11.89 – 15.81 |
|  | Tuesday | 222 | 17.98 | 15.87 – 20.23 |
|  | Wednesday | 239 | 19.35 | 17.18 – 21.67 |
|  | Thursday | 220 | 17.81 | 15.72 – 20.06 |
|  | Friday | 212 | 17.17 | 15.10 – 19.39 |
|  | Saturday | 99 | 8.02 | 6.56 – 9.67 |
|  | No data | 31 | 2.51 | 1.71 – 3.54 |
| Mineral |  |  |  |  |
|  | Coal | 956 | 77.41 | 74.97 – 79.71 |
|  | Gold | 223 | 18.06 | 15.95 – 20.32 |
|  | Emerald | 17 | 1.38 | 0.80 – 2.19 |
|  | Phosphoric rock | 6 | 0.49 | 0.17 – 1.05 |
|  | Limestone | 5 | 0.40 | 0.13 – 0.94 |
|  | Sand | 5 | 0.40 | 0.13 – 0.94 |
|  | Construction materials | 4 | 0.32 | 0.08 – 0.83 |
|  | Hydrocarbon | 2 | 0.16 | 0.02 – 0.58 |
|  | Sulfur | 2 | 0.16 | 0.02 – 0.58 |
|  | Gravel | 1 | 0.08 | 0.00 – 0.45 |
|  | Drag material | 1 | 0.08 | 0.00 – 0.45 |
|  | Copper | 1 | 0.08 | 0.00 – 0.45 |
|  | Salt | 1 | 0.08 | 0.00 – 0.45 |
|  | No data | 11 | 0.89 | 0.45 – 1.59 |
| Legal status |  |  |  |  |
| Legal | 828 | 67.04 | 64.34 – 9.66 |
| Illegal | 336 | 27.21 | 24.74 -29.78 |
| No data | 71 | 5.75 | 4.51 – 7.20 |

With regard to the legal status of the mines where emergencies occurred, the majority were legal. Nevertheless, it is of interest that 56.50% of gold, 52.94% of construction material, 33.33% of emerald, and 20.71% of coal mines were not legal. In terms of the type of emergency in illegal mines, 57.14% were from slope instability, 48.15% from flood, 34.32% from polluted air, and 31.80% from collapses. The relative frequency of injuries and fatalities was greater at illegal mines than at legal mines (*p*<0.05, Mann-Whitney test).

Table 3 shows the types of emergencies according to the mineral extracted from the mines. Most notably, all types of emergencies were more frequent in coal and gold mines. Collapses were most frequent (33.36%), followed by polluted air (oxygen-deficient atmosphere from oxygen being displaced by methane and carbon monoxide) (13.68%), explosions (most of which were associated with methane and/or dust in coal mines) (12.55%), mechanical failure (9.64%), and fires (7.94%). Unstable slopes were important in alluvial gold mining, and especially in illegal extraction.

**Table 3.**
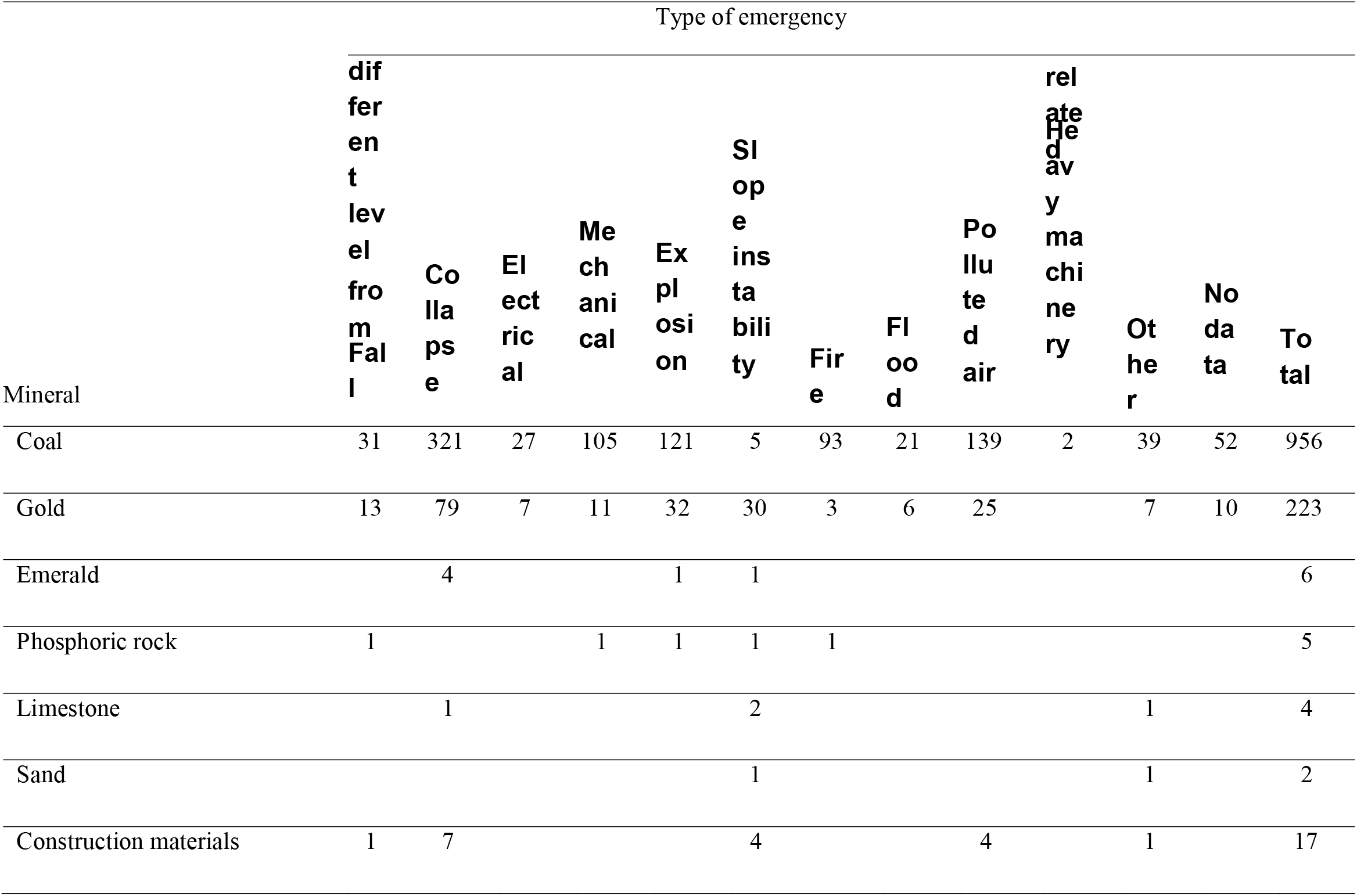
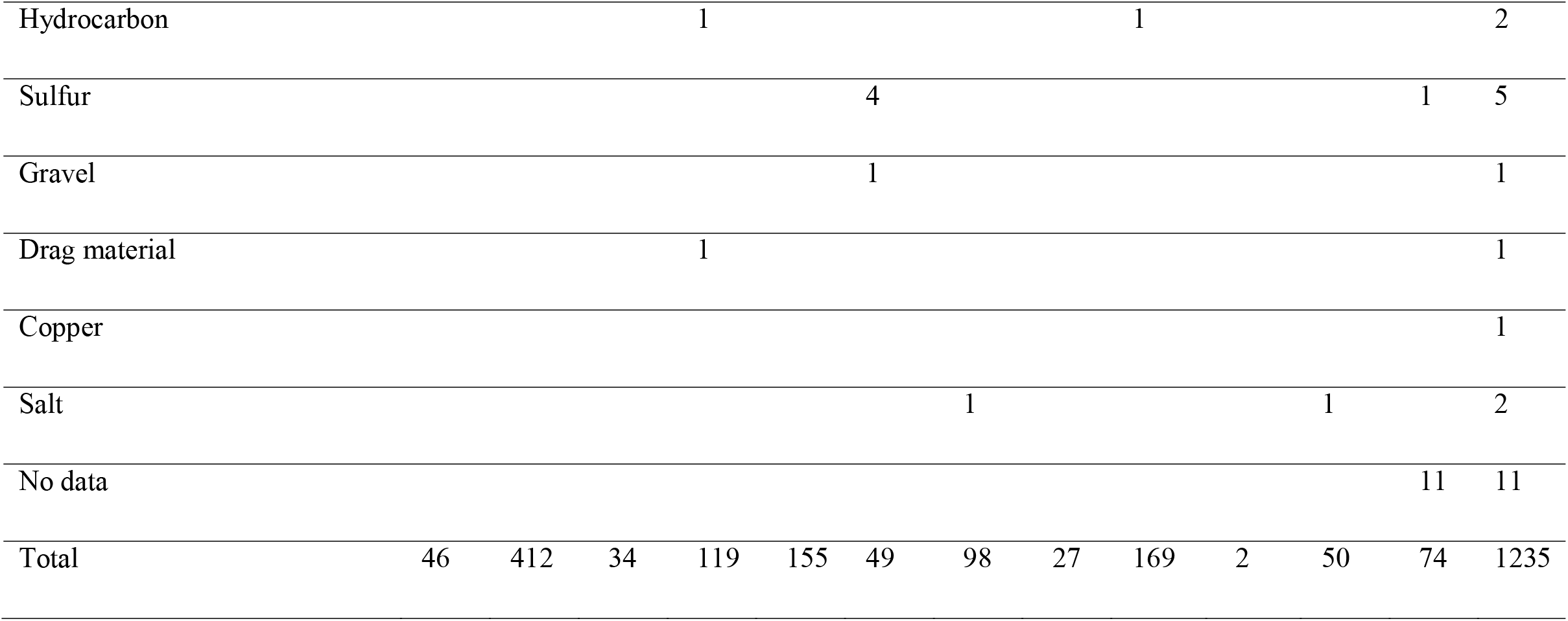
Occurrence of mining emergencies and extracted minerals in Colombia (2005-2018).

## DISCUSSION

The findings from this study suggest a high occurrence of mining emergencies in Colombia, with no clear decreasing trend over time. This is related with the increase in the amount of mines in Colombia as a result of the mining-energy boom over the past two decades ^6^. At first glance, the fact that there were more fatalities than injuries may seem contradictory. One explanation is that the data that was analyzed corresponded exclusively to mining emergencies; our study did not include occupational injuries occurred in mining context non-related with emergencies. Nevertheless, the data’s deviation from the distribution described by Benford suggests an underreporting of cases ^11^.

This type of result cannot be separated from other findings related with occupational health in Colombian mines. The concentration of suspended particles in air is a good indicator of industrial safety in coal mines. In the case of open-pit mines, which by definition have a lower concentration than underground mines, the findings indicate wide variability in Colombia, from complying with international standards to very high values such as average annual PM_10_ concentrations over 70 µg/m^3^ ^12^. In underground coal mining, an association has been reported between levels of polluted air and pneumoconiosis, which has reached a prevalence of as much as 35.9% for those with more than 10 years working in mining ^13^. Spirometric signs, symptoms, and findings also suggest respiratory problems ^14^. The coal mining costs that are associated with moral, legal, and economic responsibilities are greater than the economic benefits generated by extracting and exporting the mineral ^15^.

Gold mining has been related with serious environmental and social problems, including problems for human health, especially from the extended use of mercury that is pervasive in many regions of the country ^16^. Various effects have been seen in diverse populations of workers and the general community ^17^. With regard to underground gold mining, a lack of control over the use of mercury has led to excessively high levels in urban air, which have been registered as the highest worldwide, with up to 1 million ng/m^3^ inside gold shops ^18^. With regard to emerald mining, this has been strongly linked with violence in Colombia ^19^, which has impeded the study of safety and health conditions in those mines. A relationship with violence has also been seen in illegal gold mining ^20^.

When considering the findings presented herein, limitations due to the data that was used should be taken into account. Although official records were used from the institution responsible for addressing mining emergencies in Colombia, the evaluation of the data’s quality and the fact that there were more fatalities than injuries strongly indicate underreporting, as was also observed during the H1N1 influenza epidemic ^21^. The evidence presented in this study suggests that the majority of unreported emergencies would have been related with illegal mining, the extraction of gold, or problems with slope stability.

## CONCLUSIONS

The occurrence of mining emergencies in Colombia is high, and they affect a significant number of workers. A joint effort is needed among governmental agencies, mine owners, companies that provide employer’s liability insurance, and academic institutions in order to bring about cultural changes that prevent mining accidents and promote legal mining and compliance with occupational health and safety laws, as well as environmental protection. Mining rescue will continue to pose a challenge as long as economies are based on the extraction of materials. It is highly advisable for the Colombian mining authority to design and manage a system, which enables it to consolidate data on all serious and deadly accidents, as well as the causality analysis results of all investigations conducted. Future efforts should prioritize training and research to control the principal mining risks, including underground mine ventilation, geomechanics, hydrogeology, control of explosions associated with methane and coal dust, and the evaluation of risks and hazards.

## Data Availability

Data used in this manuscript are public.

## ACKNOWLEDGES

All authors contributed in the study (data collection and/or analysis), and participated in the writing of the final manuscript. This study is part of the master thesis of GCG and was supported by the authors.

## CONFLICT OF INTERESTS

None

## Notes

### Competing Interest Statement

The authors have declared no competing interest.

### Funding Statement

The study was supoported by the authors

### Author Declarations

The study was approved by the Ethics Committee (Universidad del Rosario, Colombia), and it is part of the GCG master's thesis.

